# Implementation, interrupted: Identifying and leveraging factors that sustain after a program interruption

**DOI:** 10.1101/2021.09.23.21263590

**Authors:** Rachel Hennein, Joseph Ggita, Bashir Ssuna, Donna Shelley, Ann R. Akiteng, J. Lucian Davis, Achilles Katamba, Mari Armstrong-Hough

**Author notes:** **Corresponding author:** Mari Armstrong-Hough.

## Abstract

**Background:** Many implementation efforts experience interruptions, especially in settings with developing health systems. Approaches for evaluating interruptions are needed to inform targeted re-implementation strategies.

**Methods:** This study took place in two public health centers with tuberculosis (TB) units in Uganda that previously implemented diabetes mellitus (DM) screening in 2017. In 2019, we conducted interviews with clinic staff to determine current DM practices. We mapped themes identified in the interviews to a Social Ecological Model with three levels: outer setting, inner setting, and individuals.

**Results:** We conducted nine interviews with clinic staff. Respondents explained that DM screening ceased due to disruptions in the supply chain for glucose test strips. This outer setting interruption had cascading effects on the inner setting and individuals. The lack of screening supplies limited the staff’s opportunities to perform DM screening within the inner setting level, which was associated with diminished self-efficacy within the individual level. However, culture, compatibility and individual beliefs about DM screening sustained throughout the interruption.

**Conclusions:** We identified factors that diminished and sustained within and between ecological levels during a program interruption. Using this approach, other programs facing interruptions can identify factors and cascading effects of the interruption to target them for re-implementation.

## Introduction

Implementation science and theories of improvement have been used to inform and assess interventions across many contexts (1). The Consolidated Framework for Implementation Research (CFIR) is among the most influential of these frameworks, and has been applied to understand implementation of programs ranging from evidence-based tuberculosis (TB) practices in Indonesia to child and maternal health outcomes in Kenyan hospitals (2-4). The CFIR was created by the Veteran Affairs (VA) Healthcare System to identify constructs that impact the successful implementation of an intervention within a series of key domains, including the outer setting, inner setting, characteristics of individuals involved, intervention characteristics, and process (5).

While the CFIR has been used to optimize the initial adoption and progress of interventions during the study period, it has not been used to scrutinize the long-term sustainability of implementation efforts when outer setting resources or characteristics vary over time (6). Variation in outer setting resources or characteristics is common in many low- and middle-income countries. For example, variation in availability of essential medicines and screening and management resources have impeded sustainable care for HIV, TB, reproductive health, and non-communicable diseases (7-12). Despite these well-reported outer setting barriers, no studies have applied the CFIR to analyze how variation in such settings may relate to program interruptions, or applied the CFIR to identify barriers and facilitators to re-implementation following significant interruptions.

Thus, there is critical need for an implementation science of interruption: guidance for how to prevent or mitigate interruptions, categorize their sources and targets, and design re-implementation strategies that build on the elements that tend to persist in the face of interruption. To respond to this knowledge gap, we carried out a qualitative case study to examine how the elements of an evidence-based intervention to screen TB patients for diabetes mellitus (DM) in Kampala, Uganda sustained, evolved, or suspended after the initial implementation period in the context of variation in key resources such as essential medicines and diagnostic materials. The World Health Organization (WHO) recommends routine screening and treatment of DM among patients presenting with TB (13) in response to a growing body of evidence that suggests a bidirectional relationship between DM and TB (14). DM has been observed to increase the progression from latent to active TB three-fold and increase adverse treatment outcomes for active TB fivefold (15, 16). However, patients presenting at TB units with hyperglycemia in low-resource settings are rarely diagnosed or counseled on controlling their blood sugar levels: half of Ugandans living with DM and 90% living with pre-DM are left undiagnosed (17).

## Materials & Methods

We conducted qualitative interviews with clinic staff using the CFIR to identify factors that persisted and factors that diminished in the midst of a program interruption. By reorganizing these CFIR-derived factors into an ecological model explicitly, we propose an approach to guide assessments of intervention interruptions and inform re-implementation strategies.

### Conceptual framework

The implementation science agenda has prioritized the identification of explanatory mechanisms that describe how the context in which an intervention is being implemented impacts sustainability (18). One way to categorize these mechanisms is through the use of Ecological Theory, which contextualizes an individual’s functioning within the context of their immediate social networks, setting, and community (19). This theory has been applied to health programs, whereby the program functions within the context of its practitioners, practice setting, public policy, and population (6). Factors at each ecological level can change over time and impact the compatibility and sustainability of the program (5, 6). Similarly, the CFIR is a multilevel framework that posits interaction among multiple domains (outer setting, inner setting, process, individual characteristics, and intervention characteristics) (20), though these levels are not explicitly nested. Thus, we sought to nest CFIR constructs within an ecological model to conceptualize how implementation factors interact between and within ecological levels during a program interruption. By situating the CFIR more explicitly within an ecological model, we aimed to understand the cascading effects and mechanisms of program interruptions at each ecological level to target them for re-implementation.

### Study setting

This case study took place in two public health centers with high-volume TB units in Kampala, Uganda. In 2017, the estimated incidence of TB in Uganda was 201 per 100,000 people, and the TB mortality rate was 26 per 100,000 (21). Public clinics in Uganda provide free healthcare to patients diagnosed with TB as long as supplies are available, whereas private clinics charge patients (22). Yet the TB treatment success rate in Uganda was 77% in 2016 (21), below the WHO target minimum success rate of 90% (23, 24).

### Initial implementation

Both health facilities had previously participated in a public health program of integrated TB-DM screening and evaluation, which ended in December 2017. This pilot program equipped staff in TB units with implementation strategies, namely training, screening algorithms to support clinical decision-making, glucometers, test strips, and TB treatment registers with entries for random blood glucose and fasting blood glucose readings. However, the inconsistent supply chain in the outer setting led to inconsistent access to key materials in the clinics after the initial implementation, such as glucose test strips and functional glucometers. Consequently, delivery of DM screening in TB clinics declined.

### Re-implementation study

In June 2019, independent of the initial pilot program, we sought to assess the implementation climate for newly introducing DM care into these two TB clinics. Neither clinic had regularly offered DM evaluation since shortly after the previous pilot ended, 18 months prior. However, during the course of ethnographic observation of the clinics, we found that many remnants of the previous DM screening program persisted, including laminated DM screening algorithms, equipment, and patient education materials. Thus, we conducted key informant interviews with health workers to determine the *re-implementation* climate for integrating DM screening into the TB clinic. We sought to identify aspects of the original DM screening implementation that persisted and diminished during the interruption to create a targeted re-implementation strategy. The Consolidated Criteria for Reporting Qualitative Research (25) guided study reporting.

### Sampling and study participants

We purposively sampled health workers and administrators from the two public health centers in August 2019. First, a male Ugandan researcher on the team (JG) introduced a female non-Ugandan researcher (RH) to the health workers at clinic sites. RH spent six weeks conducting ethnographic observation and meeting the health workers at the clinic sites to establish a relationship and share reasons for the research. Then, participants were recruited in person based on their involvement with delivery of care in the TB unit. Participants were invited to participate in key informant interviews about barriers and facilitators to the integration of DM screening into routine TB evaluation and treatment initiation.

### Data collection

We created a semi-structured interview guide using the CFIR to assess factors related to the sustainability of TB-DM care integration (5). First, we included questions from the “outer setting,” “inner setting,” and “characteristics of individuals” CFIR domains to identify factors that impact program sustainability at each of these ecological levels. As our study was designed to identify ecological factors that facilitate and mitigate sustainability and we did not anticipate a strong institutional memory of the previous intervention, we excluded the “process” and “intervention characteristics” domains initially. However, as participants discussed their experiences with the previous TB-DM program and provided recommendations for improvement, we employed probing questions related to these domains. For example, after participants suggested that the unstable supply chain hindered their ability to continue DM screening, which is related to the “outer setting” domain, we asked probing questions about protocols for dealing with stockouts, which is related to the “planning” construct within the “process” domain.

After creating the questions, we iteratively trialed the interview guide with a Ugandan researcher who is an expert in conducting qualitative interviews in this setting (JG). Through local pre-testing, we adapted the interview questions to ensure conceptual equivalence in Uganda.

All interviews were conducted at the clinic sites in English by a doctoral student with six years of qualitative interview experience in East Africa (RH). Interviews lasted from 20 to 66 minutes and field notes were made after each interview. Interviews were recorded on an encrypted smart phone. Interviews were de-identified, professionally transcribed, reviewed and revised for accuracy, and uploaded into ATLAS.ti 8. All recordings were deleted.

### Analysis

Transcripts were double coded by a non-Ugandan researcher (RH) and the code structure was reviewed and validated by other members of the study team (MAH, JG) in two cycles using a hybrid, abductive approach (26). First, using the accepted CFIR inclusion and exclusion criteria for each construct (27), we applied codes to passages that reflected relevant CFIR constructs and subconstructs using a pre-existing code tree. To emphasize that an implementation has already taken place, we modified the CFIR constructs “readiness for implementation” to “readiness for re-implementation” and “implementation climate” to “re-implementation climate.” After this first round of directed coding, we carried out an inductive content analysis, adding novel codes to the pre-populated code tree to represent themes that emerged from the transcripts but were not strictly contained by CFIR constructs. We organized all themes by CFIR domain. We applied codes independently and allowed for co-occurrence of multiple codes.

After coding, we used this combined directed and thematic content analysis to nest CFIR constructs within the ecological model. To do this, we mapped the established CFIR domains, constructs, and subconstructs to an ecological model (Figure 1). The “outer setting”, “inner setting”, and “individuals” CFIR domains each represent their own ecological level surrounding the program sustainability. Inherent to the program’s sustainability are the program’s characteristics and process of implementation, which are related to the “intervention characteristics” and “process” CFIR domains, respectively. These domains include constructs that are situated across various ecological levels. For example, the “engaging” construct within the “process” domain is defined as involving the relevant personnel in the implementation, which can be mapped to the health workers in the “individuals” ecological level or National Medical Stores personnel in the “outer setting” ecological level. Thus, we included the “process” and “intervention characteristics” domains within the ecological level pertinent to the specific factor. We then adapted this model to create a hybrid CFIR-Ecological Model to assess contributors to the sustainability of the TB-DM program. Lastly, we identified relationships between factors at different ecological levels to better understand the mechanisms and cascading effects that impact program sustainability to target them for re-implementation (28). Data saturation was defined as the point at which no new themes emerged from the data (29).

**Figure 1.**
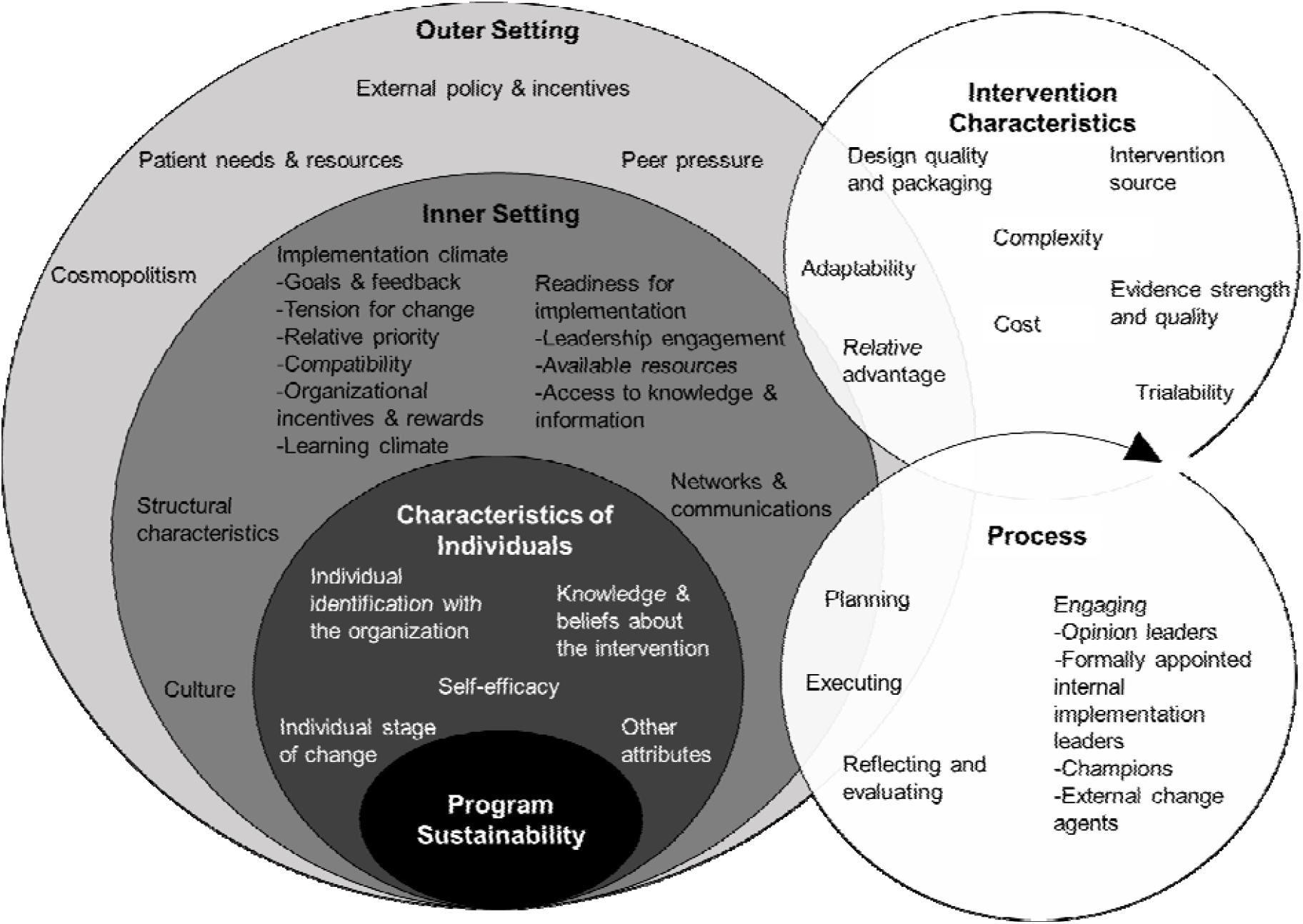
Hybrid CFIR-Ecological Model to systematically evaluate an intervention interruption. The outer setting, inner setting, and characteristics of individuals CFIR domains are each mapped to their appropriate ecological levels. CFIR constructs for each of these domains are included within the corresponding ecological level. The intervention characteristics and process CFIR domains and associated constructs span multiple ecological levels and are sorted to the appropriate ecological level based on the specific factor.

### Human subjects and ethics approval

Each participant provided verbal consent. The study was approved by the ethics review board of the Makerere University School of Public Health and Yale University Human Investigation Committee.

## Results

### Sample

Nine of nine staff members across the two public-sector TB clinics agreed to participate in the interviews (Table 1). All key informants worked at a participating clinic during the original TB-DM implementation in 2016 and continued to work in a public facility in August 2019. Their median years of experience in their current role was 3 years (range: 2 weeks, 8 years) and median years of experience in the health field overall was 8 years (range: 3 years, 20 years). Most (7, 78%) were female. As all staff members across the two public-sector TB clinics participated and no new themes emerged from the data after seven interviews, sample saturation and data saturation were met, respectively.

**Table 1.** Participant characteristics.

| **Characteristic** | **Health Center** |
| --- | --- |
| Gender, N (%) female | 7 (78) |
| Role |  |
| TB community health worker, N (%) | 4 (44) |
| TB focal person, N (%) | 1 (1) |
| TB clinician, N (%) | 1 (1) |
| TB nurse, N (%) | 1 (1) |
| Supervising nurse, N (%) | 1 (1) |
| Administrator, N (%) | 1 (1) |
| Years of experience in current role, Median (range) | 3 (2 weeks, 8 years) |
| Total years of experience in the health field, Median (range) | 8 (3 years, 20 years) |

**Abbreviations:** TB=tuberculosis

Staff members explained that the delivery of DM screening in the TB clinic was interrupted by changes in the outer setting, particularly the supply chain for glucose test strips. They expressed confidence that without disruption of necessary materials for evaluation of DM, screening would have persisted even without other ongoing support. Yet, once these materials were not consistently available, even parts of the DM screening program that did not directly rely on blood glucose testing ceased. Unpredictable supplies of materials from the National Medical Stores eventually disrupted all activities related to screening for DM in the TB units, so that even during periods when test strips were available they were no longer routinely used.

Content analysis identified one factor that was diminished and two factors that were sustained from the outer setting domain (Table 2). From the inner setting domain, two factors were diminished while three factors were sustained (Table 3). From the characteristics of individuals domain, one factor was diminished and one factor was sustained (Table 4). The CFIR domain, constructs, descriptions, and representative quotes for each factor are presented in Tables 2-4. We mapped each of these factor-domain pairs to their respective ecological level in the CFIR-Ecological Model depicted in Figure 1. Our adapted model describing the erosion or persistence of key factors influencing implementation of DM screening in the face of program interruption is represented in Figure 2.

**Table 2.**
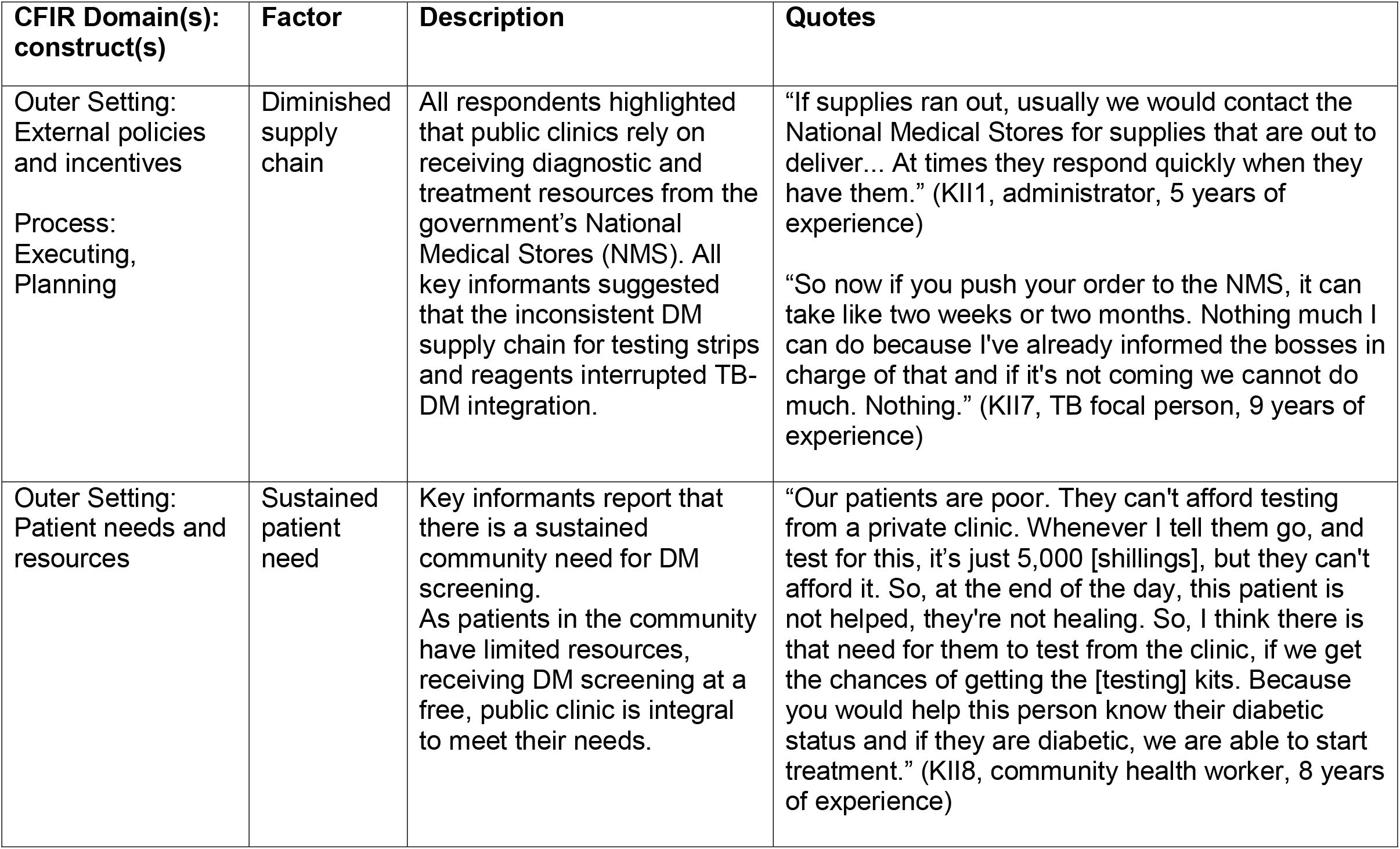
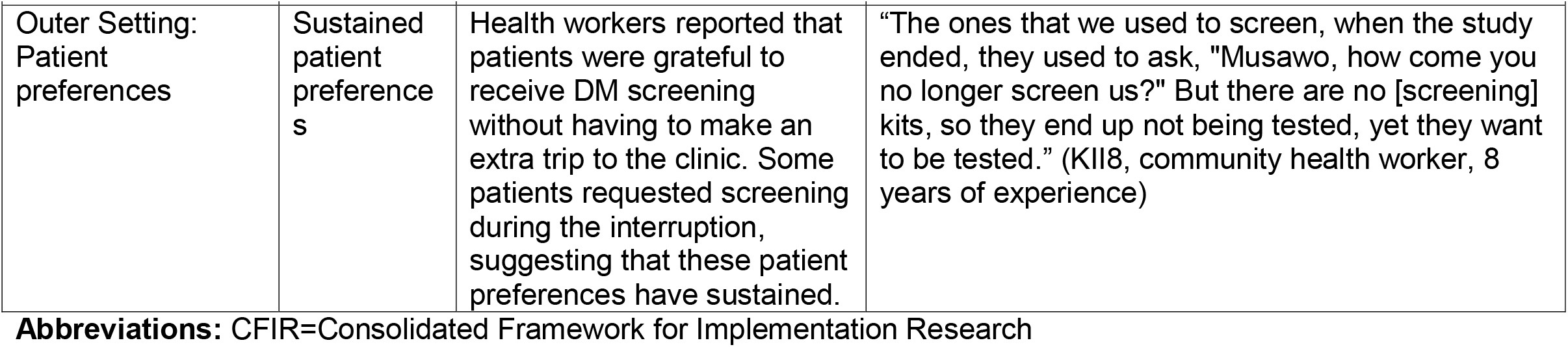
Outer setting factors that were sustained or diminished during study interruption.

**Table 3.** Inner setting factors that were sustained or diminished during study interruption.

| **CFIR Domain(s): construct(s)** | **Factor** | **Description** | **Quotes** |
| --- | --- | --- | --- |
| Inner setting: Culture, Networks and communications, Re-implementation climate<br>Process: Engaging<br>Intervention Characteristics: Complexity | Sustained compatibility with culture and workflow | Health workers reported that their clinic's collective goal is to improve community health. All respondents emphasized that DM screening is vital for community health and is therefore compatible with this goal. Respondents detailed how DM screening remains to be compatible with the TB workflow and integration would not add complexity. | "We are all aiming at achieving saving life. The bond between us, regardless of other things, is that we are all aiming at saving life especially as practitioners of TB and HIV. So when I get a client, it's not my client. It is for all of us." (KII7, TB focal person, 9 years of experience)<br>"I think it would be best practice when the patient has been confirmed with TB after getting the results from the lab, and it's suggesting TB, that same time, we first take off the blood to check the sugars. On the first day and knowing that is having tuberculosis, we also screen for diabetes as well in the same day. So when the sugar levels are high on that day, you request the patient to come back the following morning for fasting blood sugar. So if you confirm that still it's high, then you forward to the doctor, and prescribes the right drugs. Then he starts medication, alongside with TB treatment." (KII3, community health worker, 14 years of experience) |
| Inner Setting: Re-implementation climate, Readiness for re-implementation<br>Intervention Characteristics: Evidence strength and quality | Sustained prioritization of expanding DM screening by staff and leaders | Health workers highlighted that the clinic's high prioritization of DM screening by clinic staff and leaders sustained throughout the interruption. Many respondents proposed substantial expansion | "DM screening is very positive for the client's sake and also for the health workers because when you diagnose the DM in a TB client that it is good because both are deadly diseases" (KII2, Health Facility In-Charge, 20 years of experience)<br>"If we could move with a [screening] machine and test contacts also in their household, you can do it best like as you do for HIV and TB. I know we shall have results, because people down there in the community are very |

| CFIR Domain(s):<br>construct(s) | Factor | Description | Quotes |
| --- | --- | --- | --- |
|  |  | of screening efforts to screen patients and their families in their homes. | sick and they don't mind. So if we can diagnose them early now, I think it can be of good help to them." (KII5, community health worker, 8 years of experience) |
| Inner Setting:<br>Readiness for re-implementation<br>Process:<br>Executing,<br>Planning | Diminished access to screening supplies and unclear stockout protocols | The interruption in the outer setting's DM supply chain inhibited the clinic's procurement of screening supplies. There was a lack of consensus among respondents about what to do when supplies ran out. | "We need to be supplied with each and everything which is used in [DM] screening because the government doesn't provide the things... The government is not supplying enough." (KII9, community health worker, 3 years of experience)<br>"At the moment we don't have anything. We don't have the equipment. We used to have one glucometer back then. Right now it is there but of course it's not functional. We are not using it." (KII3, community health worker, 14 years of experience) |
| Inner setting:<br>Readiness for re-implementation | Diminished team learning | The lack of screening supplies did not equip the clinic to conduct ongoing training and clinical practice related to DM screening, diminishing team learning. | "Getting a training for the staff, it makes it better because it is a reminder for the staff of what you're supposed to do for the patient" (KII6, clinician, 7 years of experience)<br>"Even if you had a training before but you didn't practice, so that will be old. So you really need constant trainings so that you're up to date." (KII4, nurse, 6 years of experience) |
| Inner Setting:<br>Readiness for re-implementation | Sustained internal resources | Respondents expressed that they still have the physical space, workforce, and available time to carry out DM screening. | "We are enough. In the TB unit, now we are full with the focal person, and another nurse, and then the two of us. So we can do it best." (KII5, community health worker, 8 years of experience)<br>"We have two rooms. In the second room is where we do |
|  |  |  | HIV testing and DM screening as well. That's where we used to carry it all. So we have the space." (KII3, community health worker, 14 years of experience) |

**Abbreviations:** CFIR=Consolidated Framework for Implementation Research

**Table 4.** Characteristics of individuals that were sustained or diminished during study interruption.

| **CFIR Domain(s): construct(s)** | **Factor** | **Description** | **Quotes** |
| --- | --- | --- | --- |
| Individuals: Knowledge and beliefs about the intervention, individual identification with the organization<br>Process: Engaging<br>Intervention Characteristics: Evidence strength and quality | Sustained knowledge, beliefs, and motivation to screen for DM in the TB clinic | Health workers reported being motivated by helping patients in accordance with the clinic's collective goal of improving community health. As all respondents expressed their sustained belief that TB-DM integration is important for improving community health, they reported enjoying providing this service to patients. During the interruption, many experienced negative feelings because they were no longer able to provide this service to patients. | "In the TB clinic, when we had that training when they were just starting the program, they were telling us that one can cause the other: diabetes can cause TB and TB can cause diabetes. So, it's always good if the two are handled hand-in-hand." (KII3, community health worker, 14 years of experience)<br>"I feel bad that we are not screening DM because I know that TB can move with diabetes. We are dealing with the co-infected patients probably, they can have diabetes. If we have not screened them, they will go minus that service, so I feel bad." (KII5, community health worker, 8 years of experience) |
| Individuals: Self-efficacy<br>Process: Executing | Diminished self-efficacy to perform DM screening | Health workers expressed that the interruption hindered their ability to reinforce and improve their DM clinical skills, leading to diminished self-efficacy. | "We got some patients and were able to give care, and they are moving on well. So I think we really did a good job." (KII5, community health worker, 8 years of experience)<br>"We are taking a full year since my last doing it [DM screening]. So, somethings I may now be forgetting." (KII9, community health worker, 3 years of experience) |

**Abbreviations:** CFIR=Consolidated Framework for Implementation Research

**Figure 2.**
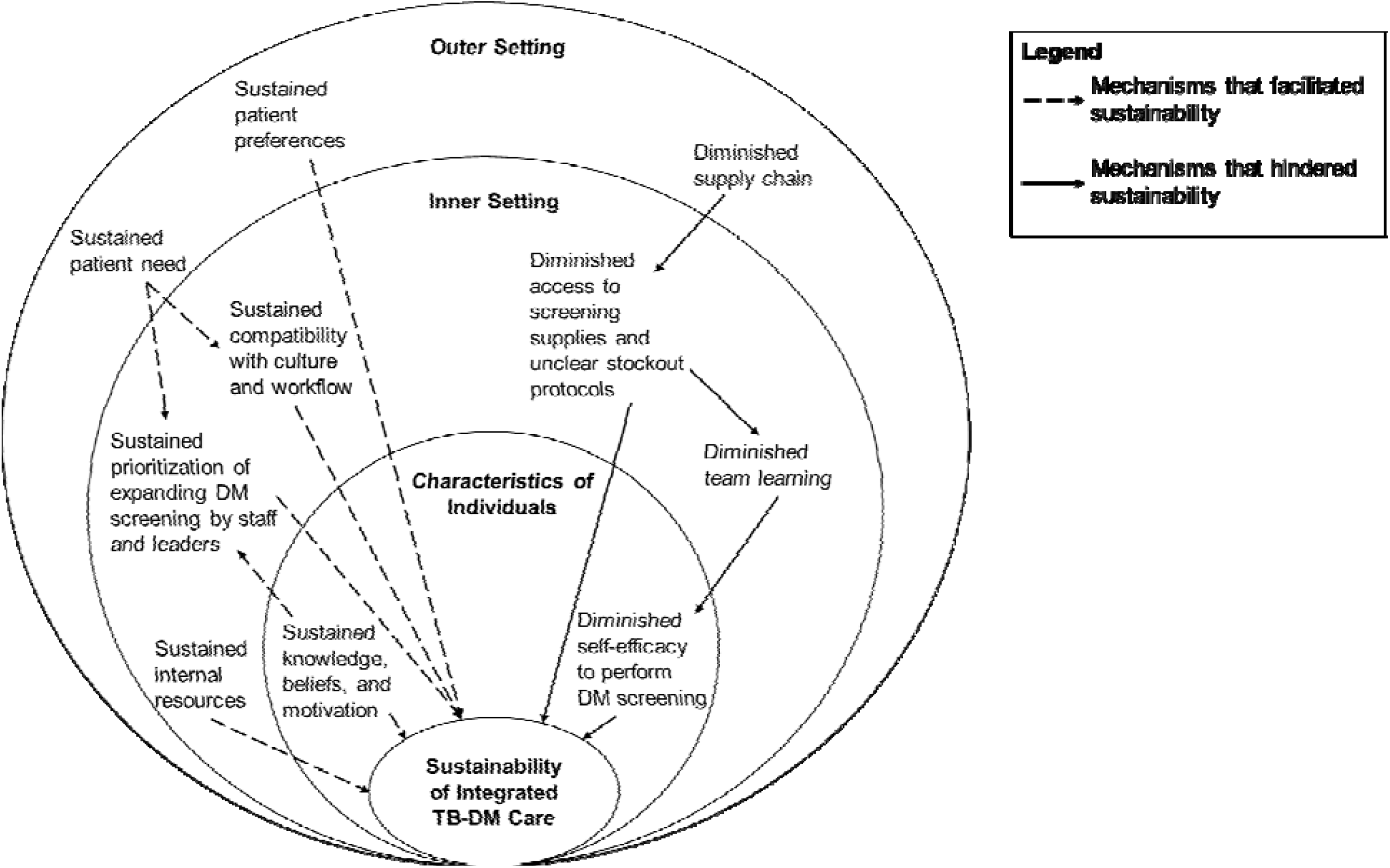
Application of the CFIR-Ecological Model to the TB-DM case study. Dashed arrows highlight the ecological dynamics contributing to the sustainability of TB-DM care during the intervention interruption. Solid arrows highlight the ecological dynamics contributing to the unsustainability of TB-DM care during the intervention interruption.

During the program interruption, key informants reported that community need and favorable patient preferences for DM screening continued (Table 2). In the inner setting, many factors facilitating DM screening persisted, including the compatibility of DM screening with the culture and workflow of the clinic, prioritization of DM screening by staff and leaders, and internal resources, such as physical space, workforce, and available time to carry out DM screening (Table 3). However, over time, the lack of DM testing kits and equipment from the outer setting diminished available screening resources and team learning in the clinic. At the level of the individual, health workers demonstrated sustained knowledge, beliefs, and motivation to screen for DM in the TB clinic long after the interruption began (Table 4). However, health workers reported that the interruption hindered their ability to reinforce and improve their DM clinical skills, leading to diminished self-efficacy.

### Cascading effects of program interruptions

There were several mechanisms by which factors from the outer setting, inner setting, and characteristics of the individuals interacted during the service interruption. First, we identified a cascading effect by which the inconsistent supply chain in the outer setting inhibited the availability of resources to screen for DM in the inner setting (Figure 2, solid arrows). Without access to screening supplies, team members could not conduct ongoing training and reinforce their skills through clinical practice. The lack of team learning in the inner setting, in turn, trickled down to impact the characteristics of the individuals. Without this sustained team learning, health workers reported diminished self-efficacy to perform DM screening.

Notably, several factors sustained during the 18-month program interruption, primarily through institutional memory of the TB-DM program’s goals and outcomes (Figure 2, dashed arrows). For example, key informants reported sustained patient need and preferences for DM screening in the outer setting. Patient needs and preferences interacted with the inner setting’s culture of altruism and reinforced their high prioritization of DM screening among staff and clinic leadership. At the individual provider level, memory and positive associations of the program led to sustained knowledge, beliefs, and motivation to screen for DM in the TB clinic. The sustained knowledge, beliefs, and motivation related to DM screening contributed to continuing high prioritization of DM screening within the inner setting at-large, even at clinics where no patients had been screened for DM for over a year. The institutional memory of the DM screening program also sustained providers’ and administrators’ perceptions of the program’s compatibility with the clinic workflow, physical space, workforce, and available time for DM screening. Providers and administrators retained positive perceptions of the intervention characteristics long after daily delivery of DM screening had ceased. The persistence of factors associated with sustainability within the outer setting, inner setting, and individual domains suggests a favorable re-implementation climate.

## Discussion

Many implementation efforts experience cycles of interruption, especially in settings with developing health systems (30, 31). Implementation programs must adapt to the dynamic context in which they function to maximize sustainable delivery (6, 32-34). However, little guidance is available for understanding or responding to interruptions when they do occur. This study adds to the literature by organizing key domains and constructs of the CFIR within an ecological model to theorize the cascading effects of outer setting instability on program sustainability. Using an empirical case study of an interruption to delivery of DM screening in Uganda, we identified implementation factors from the CFIR that persisted long after the screening program ceased, as well as implementation factors that faltered in response to changes in the outer setting. Most importantly, we described and theorized a cascading effect whereby changes in the outer setting domain first imperil constructs in the inner setting that once facilitated implementation, and through these inner setting changes eventually begin to erode individual characteristics that once facilitated implementation. By integrating the CFIR with ecological theory, we propose a framework for understanding and responding to program interruptions. This approach can be used to enhance the use of the CFIR to identify barriers and facilitators to re-implementation and target implementation factors linked to sustainability in the face of outer setting disruption.

This approach to understanding the causes, consequences, and remedies available following program interruptions is particularly relevant to other non-communicable disease programs in low-income countries. Low-income countries in sub-Saharan Africa increasingly face a double burden of disease, with high rates of infectious diseases compounded with increasing rates of non-communicable diseases (35-37). However, health systems in sub-Saharan Africa have been tailored to address infectious diseases, namely HIV, rather than non-communicable disease needs (38, 39). Variation over time in the outer setting (e.g., supply chain for essential medicines), inner setting (e.g., availability of functional glucometers), and characteristics of individuals (e.g., community health worker knowledge of diabetes) must be considered when developing, implementing, and re-implementing interventions non-communicable disease.

For example, our study found that the lack of DM screening supplies at the clinic after the pilot ended was the main outer setting barrier that led to program interruption. Outer setting shortages of technologies and tools that the WHO deems as essential for non-communicable disease care are common in sub-Saharan Africa (40, 41). Another study including 53 health centers in Uganda found that only 62% had a glucometer and 13% had urine testing strips to screen for DM. Further, only 40% had a standard blood pressure cuff and 43% had an automated blood pressure machine, which are necessary for assessing hypertension (40). Although most labs had random blood glucose capabilities (92%), only 9% could measure hemoglobin A1c and 28% could conduct a lipid profile, which are necessary for diagnosing and monitoring DM and hyperlipidemia, respectively. Lastly, all health centers reported that they experienced at least one-stock of a non-communicable disease essential medicine within the last year. While a project that provides these technologies and resources might be effective to screen and treat non-communicable diseases in the short term, studies may not be sustainable without targeting the domestic supply chains themselves.

Thus, an interruption framework for evaluating and targeting interruptions for re-implementation is critical while health systems for non-communicable diseases are further developed. In our study, we found that uncertainty in the supply chain (outer setting) hindered team learning (inner setting) and ultimately diminished the self-efficacy of practitioners (characteristics of individuals). These cascading failures could be addressed as part of a targeted re-implementation strategy by creating and disseminating tailored refresher training for key stakeholders. For example, providing targeted training to health teams in the inner setting would likely disrupt the cascading effect that decreased team learning has on individual’s self-efficacy. This training should also include the introduction of standardized protocols for dealing with stockouts in the clinic, given the lack of consensus among participants about who to contact and how to procure DM screening supplies during a stockout. We also found that the screening program in our study continued to be seen as compatible with community needs, culture, and priorities of the inner setting, as well as with motivations of practitioners. A targeted re-implementation strategy could draw on the elements that sustained during the program interruption to facilitate rapid buy-in and uptake from the community, clinic leadership, and practitioners.

### Strengths and Limitations

Our study had several strengths. First, it is strengthened by its engagement with health workers through both ethnographic and interview methods. This approach enabled triangulation, enhancing the internal validity of findings. Second, the study was informed by the CFIR from the earliest planning stages through the final analysis. We adapted the interview guide from the standard CFIR guide and carried out the directed content analysis using pre-defined CFIR construct codes. Third, we conducted an additional round of coding using inductive content analysis to generate ‘open’ codes, which ultimately generated ecological insights that form the basis of our hybrid CFIR-Ecological Model. Finally, this study was strengthened by reflecting on a two-year timeframe, which enabled key informants to comment on long-term changes.

However, this study had some limitations. Because we used purposive sampling from two public clinics in Kampala, the study setting and participants may not be representative of all public TB clinics in Kampala that participated in the earlier DM screening program. Nonetheless, many health workers report having previously worked at other public clinics in Kampala at the time of the implementation and spoke about their collective experiences. Furthermore, this study only captures the perspectives of the personnel at the inner-setting, clinic, and individual levels; thus, future studies are needed that explore interruptions in programs from the perspective of the outer setting stakeholders.

## Conclusion

We critically analyzed the long-term sustainability of a program implementation in the face of outer setting instability and used this case study together with the CFIR to produce an approach for understanding the ecological dynamics of sustainability. This hybrid approach can be used to systematically assess factors that persisted and diminished during program interruptions at each ecological level, including the outer setting, inner setting, and characteristics of individuals. Explicitly situating the CFIR within an ecological model can clarify the cascading effects of program interruptions at each ecological level and inform re-implementation strategies. Future empirical studies are needed to apply, assess, and adapt this model and further develop the study of interruption in implementation science.

## Data Availability

Data are available upon request to the corresponding author.

## Acknowledgements

We would like to acknowledge the participation of all the health workers and administrators who participated in our study and provided their insights.

